# Protocol for an Economic Evaluation Alongside the Re-Evaluating the Inhibition of Stress Erosions (E-REVISE) Trial

**DOI:** 10.1101/2025.06.23.25330120

**Authors:** Brittany Humphries, Nicole Zytaruk, Diane Heels-Ansdell, Vincent Lau, Bram Rochwerg, Robert Fowler, Yifan Yao, Deborah J Cook, Feng Xie

**Affiliations:** Department of Health Research Methods, Evidence, and Impact, McMaster University, Hamilton, Canada; Department of Medicine, McMaster University, Hamilton, Canada; Department of Critical Care Medicine, Faculty of Medicine and Dentistry, University of Alberta, Edmonton, Canada; Interdepartmental Division of Critical Care, University of Toronto, Toronto, Canada; Centre for Health Economics and Policy Analysis, McMaster University, Hamilton, Canada

**Keywords:** critical care, evidence-based medicine, health economics, gastroenterology, health services research applied methods

## Abstract

**Introduction:** Economic evaluations in healthcare can guide practice and inform policy. The objective of this paper is to present the protocol for a health economic evaluation comparing the cost-effectiveness of prophylactic treatment using pantoprazole 40 mg IV daily compared to no pantoprazole to prevent upper gastrointestinal (GI) bleed among invasively ventilated patients.

**Methods and analysis:** This is an economic evaluation conducted alongside the <u>R</u>e-<u>E</u>valuating the <u>I</u>nhibition of <u>S</u>tress <u>E</u>rosions trial. The primary outcome is the incremental cost per clinically-important upper GI bleed prevented. Secondary outcomes include the incremental cost of a patient-important upper GI bleed prevented. We will explore the incremental cost per secondary trial outcome (e.g., ventilator-associated pneumonia, *Clostridioides difficile* infection, and patient-important GI bleeding); and incremental cost per life gained. The analysis will be conducted from a Canadian public healthcare payer’s perspective over a time horizon of ICU admission to hospital discharge or death. The study protocol was developed following good practice guidelines of Canada’s Drug Agency (CDA) and the Professional Society for Health Economics and Outcomes Research (ISPOR).

**Ethics and dissemination:** The trial was approved by the Hamilton Integrated Research Ethics Board and at each participating institution; this economic evaluation is currently under review for ethics approval. Given widespread daily use of proton pump inhibitors for critically ill patients, the results of this economic evaluation will be of high relevance to patients, family members, physicians, pharmacists, policymakers and guideline developers. Integrated knowledge translation will involve periodic progress reports to collaborators. End-of-study knowledge translation will include rounds, videoconferences, abstracts and slide-decks for ICU quality councils and healthcare organizations, and open-access publications. Patient and Family Partners will co-create lay language summaries for traditional and social media to help inform all stakeholders.

## BACKGROUND

Proton pump inhibitors (PPIs) are routinely administered to critically ill patients to prevent upper gastrointestinal (GI) bleeding from stress-induced ulceration.[1] However, some studies suggest that potential adverse effects such as infection in the lungs (pneumonia) or bowels (*Clostridioides difficile*) may be more common than GI bleeding, or associated with greater morbidity, mortality, and costs.[2] As a result, recent guidelines issue conditional recommendations for PPIs in critically ill patients at high risk of bleeding based on the moderate certainty evidence.[3]

To address this gap in understanding, our team conducted the <u>R</u>e-<u>E</u>valuating the <u>I</u>nhibition of <u>S</u>tress <u>E</u>rosions (REVISE) trial [4] in which 4821 critically ill adults who were invasively mechanically ventilated were allocated to receive intravenous pantoprazole 40 mg daily or matching placebo. Clinically-important upper gastrointestinal bleeding was reduced with pantoprazole, occurring in 1.0% patients receiving pantoprazole and in 3.5% receiving placebo (hazard ratio, 0.30; 95% confidence interval [CI], 0.19 to 0.47). Death at 90 days was similar in the two groups (29.1% in pantoprazole group and 30.9% in placebo group; hazard ratio, 0.94; 95% CI, 0.85 to 1.04). Patient-important bleeding was also reduced with pantoprazole. There were no significant differences in other key secondary trial outcomes such as ventilator-associated pneumonia, *Clostridioides difficile* infection or length of hospital stay.

Health economic evaluations compare two or more healthcare interventions with respect to their costs (e.g., resource utilization) and effects (e.g., health outcomes).[5] Evaluating cost and resource use alongside the REVISE trial provides a unique opportunity to gain insight into the net effect of an intervention. The intensive care unit (ICU) is one of the most costly settings to care for patients, accounting for 20-50% of all hospital costs.[6, 7] Every dollar spent – particularly within this high-resource environment - has an opportunity cost. The cost of acid suppression in the ICU is often misunderstood because of the low acquisition cost of pantoprazole per dose.

## OBJECTIVE

The objective of this paper is to report the protocol of estimating the cost-effectiveness of prophylactic pantoprazole 40 mg IV daily compared to no pantoprazole, determined by the incremental cost to prevent a clinically-important GI bleed among invasively ventilated patients.

## METHODS

### Overview of REVISE

REVISE is an investigator-initiated, international, randomized, blinded, controlled trial (ClinicalTrials.gov NCT03374800). Patients aged > 18 years who were admitted to the ICU and expected to remain invasively ventilated beyond the calendar day after randomization were randomized to receive either placebo (0.9% sodium chloride) or pantoprazole 40 mg IV daily. Randomization was stratified by study centre and use of pre-hospital acid suppression. All other aspects of care were at the discretion of the treating team, including basic and advanced life support, enteral nutrition, and other interventions to prevent ICU-acquired complications. The primary efficacy outcome of the trial was clinically-important upper GI bleeding within 90 days and the primary safety outcome was 90-day all-cause mortality. Secondary trial outcomes included ventilator-associated pneumonia, *Clostridioides difficile* infection, and patient-important bleeding. The REVISE protocol[4] and results[8] are reported separately.

### E-REVISE design

This is an economic evaluation conducted alongside the REVISE trial (E-REVISE). The purpose of E-REVISE is to estimate the incremental cost-effectiveness of prophylactic pantoprazole 40 mg IV compared to no pantoprazole to prevent GI bleeding among mechanically ventilated patients. The primary outcome is the incremental cost per clinically-important upper GI bleed prevented. Secondary outcomes for this economic evaluation include (i) the incremental cost of a patient-important upper GI bleed prevented; (ii) an exploration of the incremental costs per difference in pre-specified secondary trial outcomes (ventilator-associated pneumonia, *Clostridioides difficile* infection, and patient-important bleeding) between the two arms; and, (iii) an exploration of the incremental cost per life gained. The analysis will be conducted from a Canadian public healthcare payer’s perspective over a time horizon of ICU admission to hospital discharge or death. The study protocol was developed following the good practice guidelines of Canada’s Drug Agency (CDA)[9] and the Professional Society for Health Economics and Outcomes Research (ISPOR).[10] A summary of the study framework is presented in **Figure 1**.

**Figure 1.**
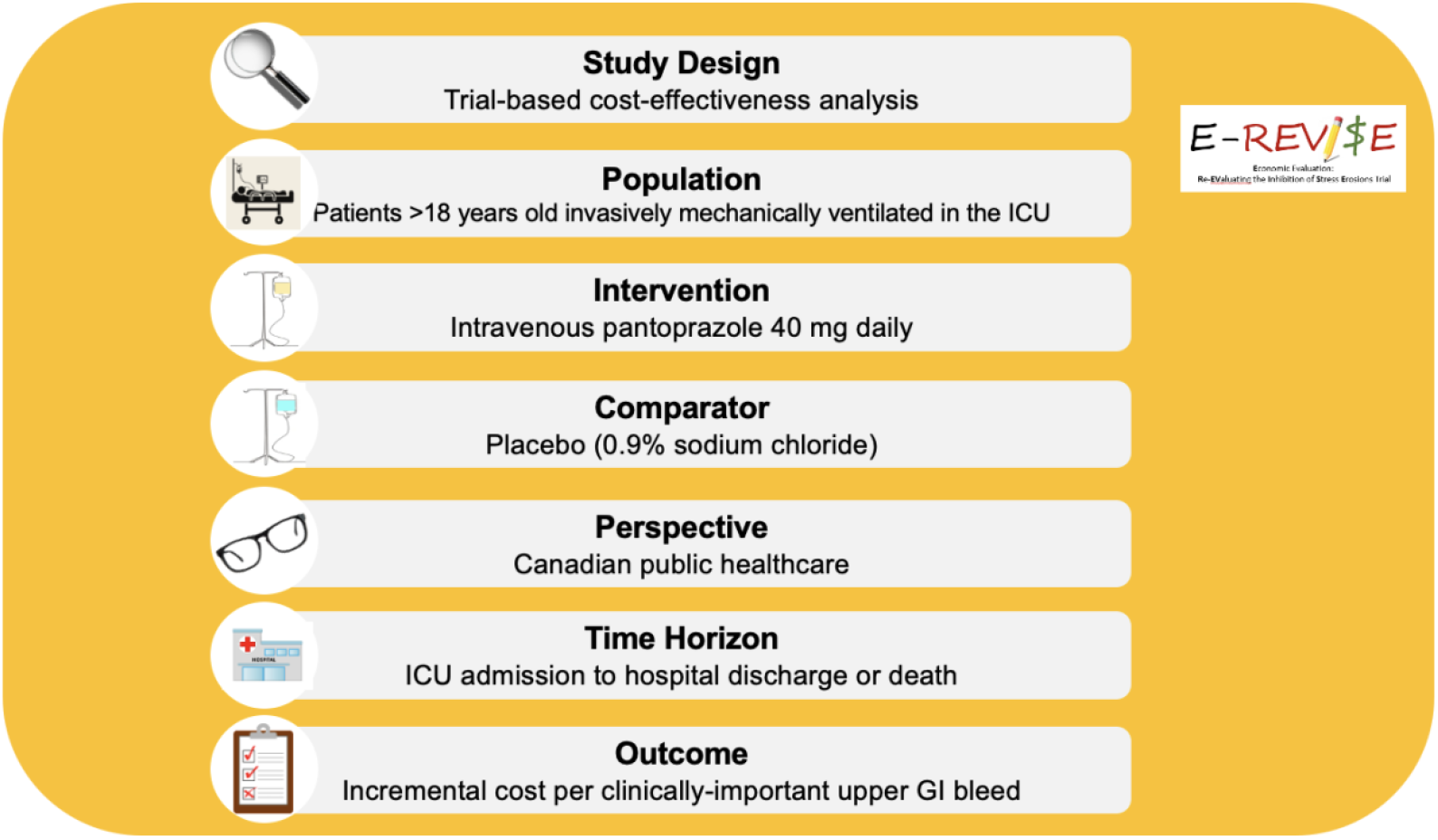
E-REVISE Framework.

### Study sample

The REVISE trial recruited 4,821 patients from 68 study centres across Australia, Brazil, Canada, United Kingdom, Kuwait, Pakistan, Saudi Arabia, and the United States. The sample size calculation, which was designed to detect a statistically significant absolute difference in the primary efficacy outcome (i.e., clinically-important upper GI bleeding), is described elsewhere.[4] This base-case analysis will focus on the 3,265 patients recruited from 42 study centres in Canada with a sensitivity analysis including the full sample.

### Data sources

We will use patient-level data collected in the REVISE trial to obtain information describing patients’ socio-demographic and clinical characteristics (e.g., age, sex, comorbidities, pre-hospital hospital acid suppression, Acute Physiology and Chronic Health Evaluation (APACHE) II score, ICU admitting diagnosis, SARS-CoV-2 status), clinical outcomes (e.g., GI bleeding, ventilator-associated pneumonia, *Clostridioides difficile* infection, death), and healthcare resource utilization (e.g., drug administration, laboratory and diagnostic tests, packed red blood cells and other blood product administration, procedures, nutrition, surgeries, advanced life support strategies, number of days in hospital and ICU). Unit costs for each resource item will be obtained from publicly available sources in Canada, such as the Ontario Drug Benefit Formulary,[11] the Schedule of Benefits for Laboratory Services,[12] and the Schedule of Benefits for Physician Services,[13] Canadian Blood Services,[14] as well as recently completed critical care health economic evaluations.[15] We will obtain additional information from wholesaler and distributors’ catalogues to address any outstanding data gaps (e.g., if data on unit cost is not available publicly). Only in-hospital costs incurred from the Canadian public healthcare payer perspective will be considered. For patients enrolled in other countries, their resource use will be estimated using the data collected in the REVISE trial. The unit costs will be based on the Canadian unit costs as a proxy.

### Statistical analyses

Descriptive statistics (mean, standard deviation, frequency, percentage) will be used to summarize patients’ baseline characteristics, clinical outcomes, and costs. The resources used will be multiplied by their unit costs and then summed to calculate individual patient costs. In the base-case analysis, the incremental cost-effectiveness ratio (ICER) will be estimated by dividing the incremental cost by the incremental effect for pantoprazole and no pantoprazole groups. Incremental effects are defined as the difference in per-patient event rates or the difference in the proportion experiencing a clinical event between groups. The primary outcome is the incremental cost per clinically-important upper GI bleed prevented. We will also compare the secondary outcomes and associated costs, as appropriate, between the two treatment arms.

All analyses will be conducted according to the intention-to-treat principle. Missing data will be reviewed to determine the amount of missing data and assess whether they are missing completely at random. If needed, an appropriate imputation method will then be selected according to the type and distribution of the missing data.[16]

All costs will be presented in 2025 Canadian dollars. No discounting will be applied due to the short time horizon (< 1 year). The analyses will be conducted using R (version 4.4.0) statistical software.

### Sensitivity analyses

To address sampling uncertainty, we will perform a probabilistic sensitivity analysis using non-parametric bootstrapping techniques to generate 95% confidence intervals (CIs).[10] One-way sensitivity analyses will also be conducted for key influential variables (e.g., cost per day cost of care in ICU) and assumptions (e.g., cost of an upper gastrointestinal bleeding event). As patient characteristics, clinical outcomes, resource use, and costs may differ across jurisdictions and outside of tightly controlled clinical trials,[17] we will conduct additional sensitivity analyses to explore how changes in values of specific variables (e.g. including costs of pantoprazole) impact ICERs. We will also conduct a sensitivity analysis by including the full trial sample using Canadian unit costs.

### Subgroup analyses

As with other REVISE trial analyses, we will conduct subgroup analyses according to: 1) use of pre-hospital acid suppression (proton pump inhibitors or histamine-2-receptor antagonists vs. no pre-hospital acid suppression); 2) APACHE II score ( ≥ 25 vs. < 25); 3) ICU admitting diagnosis (medical vs. surgical/trauma); 4) SARS-CoV-2 status (positive vs. negative); and 5) sex (female vs. male).[4]

### Patient and public involvement

In a mixed-methods study,[18] patients and family members developed a secondary REVISE trial outcome of patient-important upper gastrointestinal bleeding [19] which in the REVISE trial was also found to be reduced in those receiving prophylactic pantoprazole. Regarding the design and conduct of this economic evaluation, members of the public were not involved; however, when the economic analysis is complete, we will co-create knowledge translation interventions, involving them in the dissemination plans.

### Ethics and dissemination

All participating centres received research ethics approval before initiation by hospital, region or country, including, but not limited to – Australia: Northern Sydney Local Health District Human Research Ethics Committee and Mater Misericordiae Ltd Human Research Ethics Committee; Brazil: Comissão Nacional de Ética em Pesquisa; Canada: Hamilton Integrated Research Ethics Board; Kuwait: Ministry of Health Standing Committee for Coordination of Health and Medical Research; Pakistan: Maroof Institutional Review Board; Saudi Arabia: Ministry of National Guard Health Affairs Institutional Review Board: United Kingdom: Hampshire B Research Ethics Committee; United States: Institutional Review Board of the Nebraska Medical Centre. This economic evaluation is under review at the lead Research Ethics Board - Hamilton Integrated Research Ethics Board (REB).

We will follow the Consolidated Health Economic Evaluation Reporting Standards 2022 (CHEERS 2022) Statement[20] to report the study findings. Given universal daily PPI prescribing, the results will be of high relevance to physicians, pharmacy departments, patients and policy makers in Ontario and elsewhere. Integrated knowledge translation will involve progress reports at thrice yearly ICU and health economic fora. End-of-study knowledge translation will include rounds, videoconferences, abstracts and slide decks for ICU quality councils and healthcare organizations, and open-access publications. Patient and Family Partners will co-create lay language summaries for traditional and social media to help inform all stakeholders.

## DISCUSSION

E-REVISE is proposed in the context of a living, learning, healthcare system responsible for stewardship of scarce health resources. The REVISE trial is the largest trial undertaken comparing pantoprazole versus placebo for critically ill patients. Physicians, pharmacists and policymakers, and the general public will need to know whether the cost provides good value for their healthcare dollar. E-REVISE provides an opportunity to answer these questions and address the cost-effectiveness of pantoprazole for stress ulcer prophylaxis in ICU. A healthcare economic evaluation is critical to understand the effect of any modest differences across clinical outcomes, particularly for differences in the rare but serious *Clostridioides difficile* infections that may lead to additional resource consumption. This lifecycle approach to health technology assessment in E-REVISE is innovative in the contexts of both critical care and health economics, where attention is typically focused on new drugs or devices.[21]

### Strengths and Limitations

This health economic evaluation has certain strengths, including measurement of clinical and economic data alongside a randomized clinical trial. This offers several advantages, such as the effect of randomization to help ensure comparable baseline characteristics between groups. In addition, comprehensive collection of clinical and economic data within a trial reduces data collection time and costs, especially for data that are impractical to obtain retrospectively.[17] Timely economic data can be useful to healthcare policymakers locally and at the level of jurisdictional health services delivery (e.g., provincial in Canada) to aid budgetary and healthcare resource allocation decisions. This is especially true if the intervention being evaluated is already in current practice or is the current standard of care, such is the case with pantoprazole.

This economic evaluation also has some limitations. One limitation, which is common in trials, is that the same treatment effects and costs may not be observed in routine clinical practice. This may limit the generalizability of findings. To reduce the potential for investigator bias, particularly as the REVISE results have been published,[8] we have pre-specified the parameters for the analysis (e.g., subgroup and sensitivity analyses); furthermore, analysts conducting this health economic analysis will be blinded to study drug until the final analysis is complete. Another limitation is the short time horizon of the analysis and lack of out-patient follow-up as additional costs incurred after discharge by patients and their caregivers will not be included. While the trial recorded where patients went after discharge (e.g., acute care facility, home care, long term care), these downstream costs are out of scope of the current analysis. Only a public healthcare payer perspective was considered to align with Canadian HTA recommendations.[9] The REVISE case report forms were designed to collect key information for the randomized trial; as such, some data relevant to this health economic evaluation need to be derived from other sources (e.g., dosage of certain medications, resource use associated with pneumonia). Finally, REVISE was designed to examine the clinical effect of the prophylactic drug on upper gastrointestinal bleeding. Willingness-to-pay thresholds for this outcome have not previously been developed; in distinction to outcomes such as the incremental cost per life, life-year, or quality-adjusted life-year gained. Therefore, our results may need to be interpreted in comparison with the clinical outcomes and costs of other interventions, and by patient and health system stakeholders to assess relative value.

## CONCLUSION

E-REVISE will complement the REVISE trial with a pre-specified prospective comprehensive economic evaluation. Cost-effectiveness analyses can aid in clinical guideline development and be useful to healthcare policymakers to aid in resource allocation decisions.

## CONTRIBUTION STATEMENT

DJC, NZ, VL and FX conceived the idea; BH, DJC, YY, VL, DHA BR, RF, NZ, and FX participated in the design of the study including the analysis plan; BH drafted the manuscript; all other authors revised the manuscript and provided important intellectual content. All authors read and approve the final version of the manuscript. FX is the guarantor.

## DATA AVAILABILITY STATEMENT

The REVISE dataset will be used for secondary observational studies such as this one, addressing additional hypothesis-driven questions. Access by REVISE investigators to the REVISE dataset will follow a submitted rationale, analysis plan and approval by the Management Committee. Requests for access to the REVISE dataset by external investigators will be considered following a submitted rationale, analysis plan and approval by the Management Committee and research ethics boards as relevant. Requirements will be stipulated in a pre-specified data sharing agreement. Only de-identified trial data and standard operating procedures will be provided with data dictionaries, transferred via a secure web portal.

## LOGO

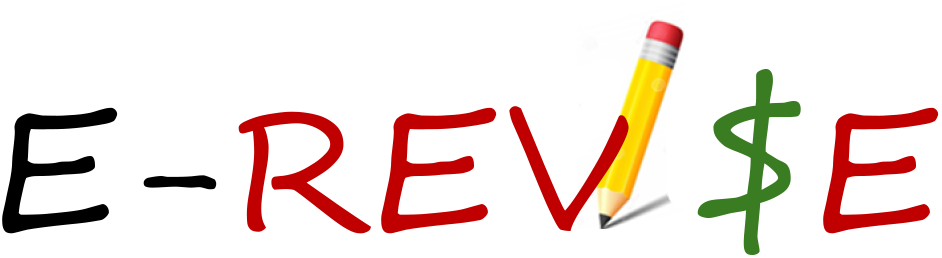

## ACKNOWLEDGEMENTS

The funding agencies had no role in the design and conduct of the study, the collection, analysis and interpretation of the data, or in the preparation, review or approval of the manuscript. D. Cook is supported by a Canada Research Chair in Critical Care Knowledge Translation from the Canadian Institutes of Health Research. B. Rochwerg is supported by a Mid-Career Award from the McMaster University Department of Medicine. R. Fowler is supported by the H. Barrie Fairley Chair in Critical Care at the University of Toronto. We appreciate discussions with Ms. C. Wallace about wholesaler and distributor catalogues for this economic evaluation. We appreciate information from Dr. K. Webert representing Canadian Blood Services. We are grateful to D. Foster for reviewing this protocol for the Canadian Critical Care Trials Group.

